# COVID-19 knowledge, risk perception and precautionary behaviour among Nigerians: A moderated mediation approach

**DOI:** 10.1101/2020.05.20.20104786

**Authors:** S.K. Iorfa, I.F.A. Ottu, R. Oguntayo, O. Ayandele, S.O. Kolawole, J.C. Gandi, A.L. Dangiwa, P.O. Olapegba

## Abstract

**Introduction:** Individuals who have knowledge of an infectious disease and also perceive the risks associated with such infectious disease tend to engage more in precautionary behaviour; however, little is known about this association as it relates to the novel Coronavirus (COVID-19). There is possibility of moderated mediation effect in the association between these variables.

**Objectives:** To examine whether risk perception determines the association between COVID-19 knowledge and precautionary behaviour among Nigerians, taking into consideration the gender differentials that may exist in the process.

**Design:** A web-based cross-sectional study.

**Setting:** Participants were recruited via social media platform, WhatsApp using google form from March 28 to April 4, 2020.

**Participants:** 1500-Nigerian (mean age =27.43, SD=9.75 with 42.7% females and 57.3% males) were recruited from 180 cities in Nigeria using snowball sampling technique. They responded to an online survey form comprising demographic questions and adapted versions of the Ebola knowledge scale, SARS risk perception scale and a precautionary behavior scale.

**Result:** Moderated mediation analysis showed that risk perception mediated the association between COVID-19 knowledge and precautionary behavior and this indirect effect was moderated by gender. Having correct knowledge of COVID-19 was linked to higher involvement in precautionary behavior through risk perception for females but not for males. COVID-19 awareness campaigns may target raising more awareness of the risks associated with the infection in order to make individuals engage more in precautionary behaviors.

**Conclusion:** Awareness campaigns and psychological intervention strategies may be particularly important at the moment, for males more than females.

## INTRODUCTION

Coronavirus disease 2019 (COVID-19) is a ravaging infectious viral disease caused by severe acute respiratory syndrome coronavirus 2 (SARS-CoV-2),[1]. Due to the rapidly increasing contagious nature of the coronavirus, which is overwhelming critical care and frontline healthcare staff and the possibility of transmission by asymptomatic carriers, governments around the world, closed their borders, announced total or partial lockdown, restrict movements, initiate social distance and facemask wearing regulations,[2, 3, 4]. The total number of infected persons worldwide as at May 15, 2020 had risen to 4,628,549 with over 308,645 deaths across 213 countries and territories of the world, [5]. Specifically, all African countries have been hit with the pandemic resulting to 79,931 infected persons and 2,640 deaths, and there are 5,450 confirmed cases and 140 deaths in Nigeria (as at 15 May 2020),[5]. Precautionary behaviour such as the extent (e.g., self-isolation, closing down of schools and work places) and mode (e.g., frequent hand washing with soap, social distancing, etc.) of human contact behaviours are identified as infection control measures which may help curtail the spread of infections,[6]. It was revealed that during periods of disease outbreaks, people who are more knowledgeable about the outbreak tend to worry more about being infected,[7], suggesting a link between knowledge and risk perception. It is worthy of note also that the trajectory of an infectious disease outbreak is often affected by the behaviour of individuals, and the behaviour is often related to individuals’ risk perception, [8,9,10]. Although there is relatively high knowledge of COVID-19 among Nigerians,[11], there is the possibility that the misunderstandings may be downplaying precautionary behaviour among Nigerians.

Therefore, it is important to investigate how COVID-19 knowledge and risk perceptions may be influencing precautionary behaviours among Nigerians, specifically if risk perception is mediating the relationship between COVID-19 knowledge and precautionary behaviours among Nigerians. In line with Gustafson’s argument that gender structures give rise to systematic gender differences in the perception of risks,[12], we also sought to investigate if gender would moderate the mediating path from COVID-19 through risk perception to precautionary behaviour. It is therefore hypothesised that COVID-19 knowledge will predict greater precautionary behaviours among Nigerians. Also, that risk perception will predict increased precautionary behaviour. Finally, it is expected that risk perception will mediate the prediction of precautionary behaviour by COVID-19 knowledge and that this effect will be stronger for female Nigerians than for male Nigerians. The conceptual model of the expected moderated mediation is shown in Figure 1 below.

**Figure 1.**
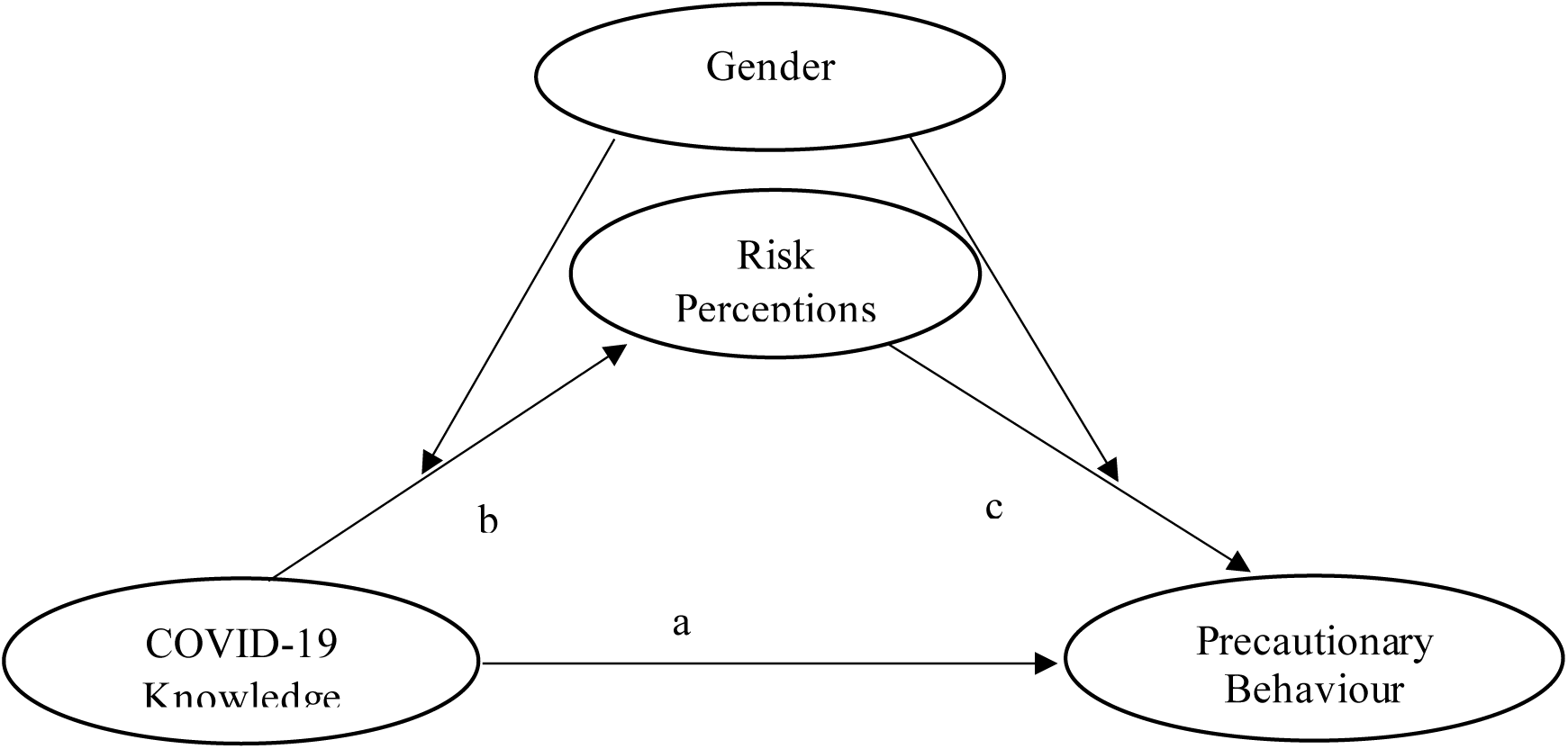
Conceptual model of moderated mediation for the effects of COVID-19 Knowledge and risk perception on precautionary behaviour

## METHODOLOGY

### Design, procedure, and sample

This was a web-based cross-sectional survey study conducted via social media (Facebook and WhatsApp) using google form from March 28 to April 4, 2020. Participants were 1554 persons (42.7% females and 57.3% males; mean age = 27.43, SD= 9.75) who responded to the survey instrument via Google Forms. They were recruited through a snowball sampling technique via social media posts to complete the online survey on COVID-19 knowledge, perceptions and precautionary behaviour using Google forms website. Participants indicated their consent to participate in the study by clicking the “next” button after reading the informed consent. Inclusion criteria included access and ability to use the internet, ability to read and write in English, willingness to sign the informed consent form and being above 15 years.

### Ethical Considerations

Ethical approval was sought from the Faculty of Social Sciences Ethical Board, University of Ibadan and participants consented for completing the questionnaires. Strict adherence to ethical provisions on confidentiality and autonomy were also observed.

### Measures

Using a pre-established Google Form, we collected data on participants socio-demographics such as; gender, age, marital status, ethnicity, educational qualification, religion and perceived financial situation.

#### Knowledge scale

Knowledge of COVID-19 was assessed using five items adapted from the Ebola knowledge scale,[13]. Respondents’ knowledge of COVID-19 is arrived at by summing correct responses across item 1, source of COVID-19, (correct = [d]), item 2, transmission of COVID-19, (correct = [a], and [b], [c] or [d]), item 3, prevention of COVID-19, (correct = [b] and [d], [f] or [h]), item 4, symptoms of COVID-19, (correct = [a], [b] and [g]), and item 5, awareness of COVID-19 fatality, (correct = [a]), generating a maximum possible score of five. The mean score and standard deviation for the sample population is calculated and scores above the norm are indicative of high knowledge of COVID-19, while scores below the norm indicate low knowledge of COVID-19.

#### Precautionary behaviour scale

Precautionary behaviour was assessed using ten adapted items,[14,15]. The ten-item scale comprises of statements dealing with actions taken in advance to protect against possible exposure to COVID-19. Sample items include: “I prefer to wash my hands pretty soon after shaking someone’s hand,” and “I am comfortable going to very crowded places (reverse scored).” Participants rated items on separate 7-point scales (1 strongly disagree; 7 strongly agree). Items 4 and 6 are reverse scored. The 10 items score are summed to obtain a composite precautionary behaviour score. A reliability coefficient (Cronbach’s alpha) of .80 was obtained in a pilot testing of the scale while the current data set yielded alpha of .75

#### Risk perceptions

To measure COVID-19 risk perceptions, the authors adapted a nine-item scale assessing SARS risk perception (e.g., “What level of threat do you think the COVID-19/Coronavirus pandemic poses to your job or studies?” and “How worried are you about contracting the Coronavirus?”),[16]. Participants rated these items on separate 7-point scales (1 = not at all likely, 7 = extremely likely). A reliability coefficient (Cronbach’s alpha) of .71 was obtained in a pilot testing of the scale while the current data set yielded .75

### Data analytic strategy

Due to the sampling of participants from different cities (six geopolitical zones), we tested the intra-class correlation coefficient (ICC) to determine the degree to which the data is dependent. The variance attributed to city of location were 0 for all variables (ICCs = 0.00), indicating that portion of variance in precautionary behaviour, COVID-19 knowledge and risk perception were independent of city (geopolitical zones). Pearson’s correlation was used to establish the relationship between the demographics and major variables of interest. For the major aims of the study, moderated mediation was carried out with the model 58 of PROCESS macro for SPSS,[17].

## Results

Correlations of the variables which were computed separately for males and females are shown in Table 1. Among females, being older (older age) was related to higher COVID-19 knowledge, and higher precautionary behaviour, but not risk perception. Higher COVID-19 knowledge was related to greater risk perception and greater precautionary behaviour. Higher risk perception was related to greater precautionary behaviour. For males, being older (older age) was related to higher COVID-19 knowledge, higher risk perception, and higher precautionary behaviour. Higher COVID-19 knowledge was related to greater precautionary behaviour, but not to risk perception. Higher risk perception was related to greater precautionary behaviour.

**Table 1.** Means, standard deviations, and intercorrelations of study variables separated by gender (n = 1551)

| Variables | 1 | 2 | 3 | 4 |
| --- | --- | --- | --- | --- |
| 1.Age | — | .09* | .04 | .10** |
| 2.COVID-19 Knowledge | .10** | — | .09* | .18** |
| 3.Risk perception | .10** | .06 | — | .23** |
| 4.Precautionary behaviour | .10** | .18** | .25** | — |
| Females (n=664) |  |  |  |  |
| Mean | 25.52 | 3.71 | 35.69 | 56.90 |
| SD | 8.22 | .82 | 10.41 | 10.69 |
| Males (n=890) |  |  |  |  |
| Mean | 28.85 | 3.71 | 35.94 | 55.80 |
| SD | 10.53 | .83 | 10.66 | 10.62 |

Note. \*\* p < 0.01; \* p < 0.05; Correlations for females are above the diagonal while correlations for males are below the diagonal

Table 2 indicated that self-isolation was the most strongly agreed upon precautionary behaviour among our present sample, followed by covering mouth when sneezing, washing hands/using hand sanitizers, avoiding crowded places, changing lifestyles, avoiding touching surfaces, and frequent testing. Face masks wearing was the least strongly agreed upon precautionary behaviour among our present sample of Nigerians.

**Table 2.**
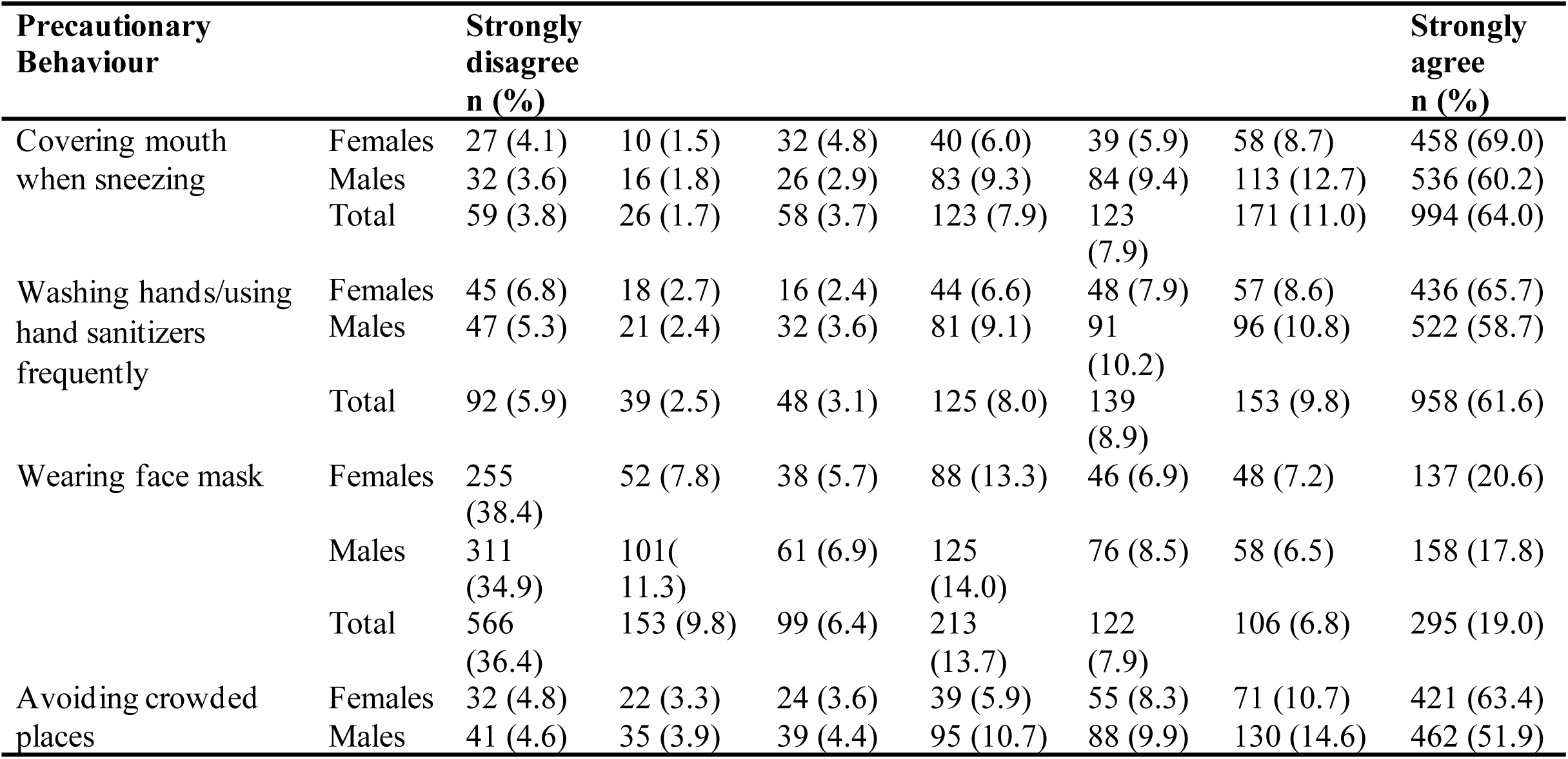
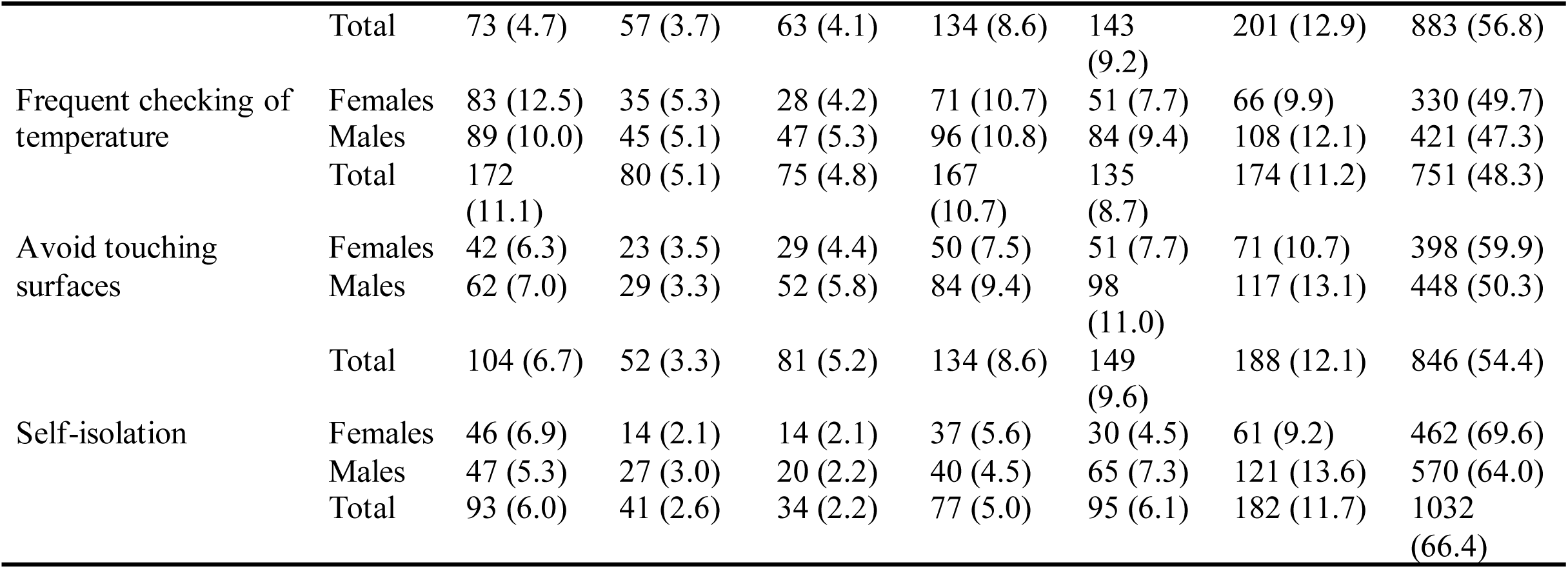
Precautionary behaviours engaged in by Nigerians

Table 3 showed that older age predicted increased risk perception. Gender did not predict risk perception. Greater COVID-19 knowledge predicted elevated levels of risk perception. Gender did not moderate the association of COVID-19 knowledge and risk perception, given that the interaction term was not significant. The predictors accounted for 1% of the variance in risk perception (R^2^ = 0.01, F (4, 1549) = 4.07, p =.003).

**Table 3.**
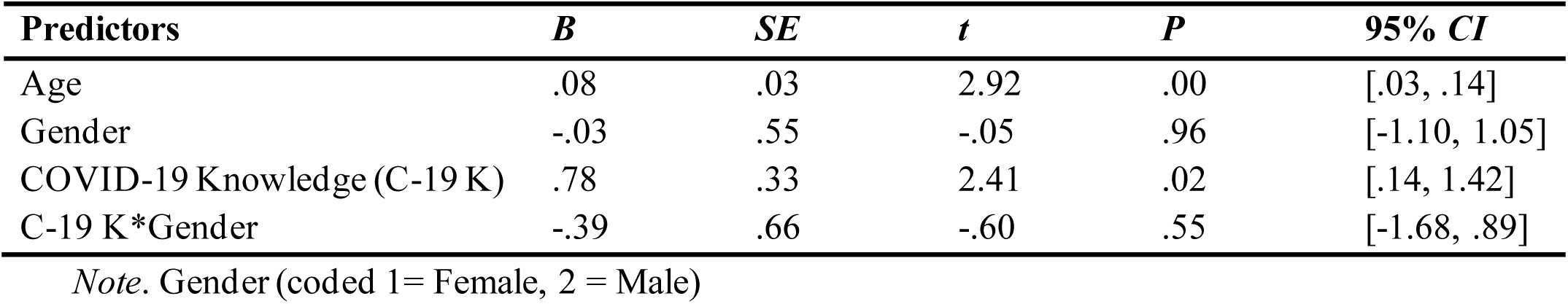
Regression results predicting risk perception by COVID-19 knowledge and gender, with age as a covariate

In Table 4, it was found that older age predicted increased precautionary behaviour. Gender also predicted precautionary behaviour suggesting that females engaged in more precautionary behaviour than their male counterparts. Greater COVID-19 knowledge predicted elevated levels of precautionary behaviour. Higher levels of risk perception predicted higher levels of involvement in precautionary behaviours. Gender did not moderate the association of risk perception and precautionary behaviour, given that the interaction term was not significant. Our hypothesis of a moderated mediation effect was supported as evidenced by a significant indirect effect of COVID-19 knowledge on precautionary behaviour through risk perception among females {B = 0.22, 95% CI = [.01, .50]}, but not males {B = 0.14, 95% CI = −.05, .37}. Note that the moderated mediation is significant when the 95% CI did not encompass zero as shown in the case for females,[18]. The predictors accounted for 9% of the variance in precautionary behaviour (R^2^ = 0.09, F (5, 1548) = 31.46, p =.00).

**Table 4.** Regression results predicting precautionary behaviour by COVID-19 knowledge, risk perception and gender, with age as a covariate

| Predictors | *B* | *SE* | *t* | *p* | 95% *CI* |
| --- | --- | --- | --- | --- | --- |
| Age | .08 | .03 | 2.82 | .01 | [.02, .13] |
| Gender | -1.42 | .53 | -2.68 | .01 | [-1.10, 1.05] |
| COVID-19 Knowledge (C-19 K) | 2.04 | .32 | 6.47 | .00 | [1.42, 2.66] |
| Risk perception | .23 | .03 | 9.25 | .00 | [.18, .28] |
| Risk perception*Gender | .01 | .05 | 2.82 | .83 | [-.09, .11] |

*Note.* Gender (coded 1= Female, 2 = Male)

## Discussions

Consistent with previous findings therefore, we postulated that perception of risk is a pathway through which knowledge and awareness of COVID-19 will influence precautionary behaviour and that this influence may be more for females than for males. We also tested the direct influences of COVID-19 knowledge, risk perception, age and gender on precautionary behaviour.

Findings showed that COVID-19 knowledge had a significant influence on precautionary behaviour. This supported our hypothesis and mirrors previous findings,[16,19]. It is logical to expect that when individuals are aware of threats, they adopt reasonable behaviours which may avert the threat from causing harm. However, sometimes people experience attitude-behaviour discrepancy which may explain human tendency to cause tragedy for the group due to inherent self-interest as can be seen in pockets of disobedience to precautionary health behaviours in the face of the pandemic.

As hypothesized, risk perception significantly predicted precautionary behaviour and this is in agreement with previous findings,[16, 20]. This implies that, perception of risk is an important variable that could inform valid and reliable precautionary behaviours and possible means of preventing a newly emerging contagious disease like covid-19. Those studies supporting the present result suggested that individual’s ability to promote precautionary behaviour largely rely on perceived risk of contracting a disease, and that risk perception is a strong predictor of precautionary behaviours. In addition, for individuals to willingly indulge in precautionary behaviours, they have to first and foremost significantly perceive the risk that such disease poses to them.

Being older predicted more precautionary behaviour, this is supported by the previous studies,[21]. These findings found that older people and those with underlying comorbid diseases took more precautionary measures compared with younger people. International and national medical agencies have put it that, older people and those with underlining disease are more vulnerable to contract the Coronavirus and may find it difficult to survive it,[22]; this may explain why elderly respondents reported engaging more in precautionary measures compared to the younger respondents.

Females reported more precautionary behaviour than males. This result is in line with the past literature where females have been consistently found to engage more in precautionary behaviour than their male counterparts,[16, 23]. This implies that, females have more tendency than males to engage in precautionary behaviours such as; washing of hands, using of hand sanitizer, nose masks, cleaning of surfaces, and having plans to visit hospital or call emergency numbers in case of any symptoms etc. It is possible that the general perceived vulnerability of females to illness,[15, 24]. This relationship between gender and precautionary behaviour may probably be that females perceive themselves as more susceptible, for example to Covid-19, than males do.

Risk perception mediated the relationship of COVID-19 knowledge and precautionary behaviour. Previous studies are similar to this result, for instance, a study conducted among Saudi and non-Saudi Arabian pilgrims in 2014 on the outbreak of Middle East respiratory syndrome coronavirus showed that overall knowledge of causative agents, the symptoms of the virus and its similarity to the disease and risk perception of the virus are associated with precautionary behaviour,[25]. The results revealed that knowledge influences precautionary behaviour through perception of risk. That is, individuals who have high knowledge but do not perceive it as a risk may not engage in precautionary behaviour.

Gender moderated the indirect path of COVID-19 knowledge to precautionary behaviour through risk perception; although, no previous study was found to have investigated if gender moderates the path way of knowledge/awareness of an infectious disease through risk perception to precautionary health behaviour; our study has contributed this knowledge and hence its availability will enhance further research in this area by other researchers. Though no direct study with similar focus and results were found, a meta-analytic study on gender differences in risk perception pointed out that gender influences perception of risk,[26]. Although, we may not be able to address the question of why gender differences exist in the pathway from knowledge through risk perception to precautionary behaviour in this study, it may be interesting to try to relate this finding to previous postulations. Owing to the offspring risk hypothesis,[27], females have a tendency to perceive greater risks than males because they are primarily care givers by nature and if one perceives more risks in the world, one will possibly be more effective at keeping safe any offspring under one’s care. Moreover, this finding corroborates a recent study which showed gender difference in risk perception of drug use between males and females, with females reporting higher levels of risk perception and therefore precautionary behaviour compared to men,[28].This result implies that risk perception serves as a path way through which COVID-19 knowledge influences precautionary behaviour and that this may be higher in females than in males.

## Data Availability

All the data involving Ethical issues such as; confidentiality, right of participants, inform consent, approval from local institutions, data analysis, questionnaire, plagiarism check and none conflict of interest are available for evaluation.

## Implication of findings

The dynamic nature of infectious disease transmission suggests that behaviour by a modest number of individuals may have a significant impact on the trajectory of an outbreak,[8-10, 29]. However, individuals may not get to take precaution if they are not aware or have the wrong knowledge about the outbreak,[30]. Therefore, in line with the findings of our study that greater COVID-19 knowledge predicted greater precautionary behaviour, and coupled with the fact that there are already myths and conspiracy theories surrounding the origin and nature of COVID-19, we recommend massive campaigns aimed at promoting correct knowledge of the COVID-19. In places where such knowledge is already influenced by conspiracy theories, it may be important to reorient the public on the real nature and origin of the disease.

In unaffected areas, true risks may be low, but due to the massive media coverage of the COVID-19, there is the possibility of elevated levels of risk perception. Therefore, the scientific community may leverage on this to explore ways to best communicate risks to individuals without unnecessarily raising panic. Individuals who perceive themselves as being at risk of contracting the COVID-19 may engage in precautionary behaviour as found in this study but may also form stereotypes and prejudices against persons perceived to be the sources of the disease outbreak as reported in our earlier findings,[11, 30]. It is therefore necessary that knowledge, realistic risks and effective precautionary behaviours be communicated through various information sources.

Consequently, individuals may need to be informed about the potential risks of infection in order to adopt the right precautionary measures,[16]. Measures to control future outbreaks should not be limited to the development of vaccines, but also adequately informing the public about the true nature (and origin of the infections) as well as risks since these have shown to be predictors of precautionary behaviour.

